# Lung adenocarcinoma WHO histological classes contain distinct immune cell profiles

**DOI:** 10.64898/2026.03.24.26348030

**Authors:** Anca Nastase, Michael Olanipekun, Elizabeth Starren, Saffron A.G. Willis-Owen, Amit Mandal, Clara Domingo-Sabugo, Deborah Morris-Rosendahl, Eric Lim, Liming Liang, Andrew G Nicholson, Miriam F. Moffatt, William O.C. Cookson

## Abstract

Lung adenocarcinoma (LUAD) is classified internationally into six histological subtypes that predict clinical outcomes. Mutation analyses identify targets but provide less prognostic information than histological appearances. Immunotherapy in LUAD is constrained by the unpredictable immune environment within tumours. We therefore characterised relationships between WHO histological classification, common mutations, and underlying transcriptomic and immune profiles in 89 LUAD cases. Mutation profiles poorly correlated with histology or survival. Global gene expression was structured into 12 modules, identifying different tumour cells and pathways within WHO subtypes. Tumour classes also held distinctive immune cell profiles. Transcripts within high-risk solid tumours indicated enrichment of CD8+ and activated CD4+ T-cells, suggesting responsivity to immunotherapy. Independently from histologic classification, 31 transcripts were strongly associated with survival and were enriched in macrophage and fibroblast derived networks. The results suggest histological subtype stratification and typing for survival-associated markers have the potential to inform clinical trials of LUAD.

## Introduction

Lung adenocarcinoma (LUAD) is the most common subtype of lung cancer worldwide and carries a high mortality. It is closely associated with smoking but is appearing with increasing frequency in never-smokers.

LUAD is classified histologically according to the 2021 World Health Organization (WHO) guidelines^1^. The classification predicts clinical prognosis, with a grading spectrum from 1 to 3 based on architectural patterns for non-mucinous adenocarcinomas. Grade 1 describes lepidic predominant with <20% high-grade patterns; Grade 2 acinar and papillary with <20% high-grade pattern; and Grade 3 includes cribriform, micropapillary and solid tumours^1,2^

A poor prognosis may be suggested by genomic analyses that identify higher tumour mutation burdens and *TP53* mutations^3,4^. Specific therapies for *EGFR* mutations are effective and although infrequent, mutations in *BRAF MET*, and *ERBB2* may also impact clinical management. Nevertheless, mutation profiles predict active proliferation and poor outcomes to a lesser degree than indicated by high-grade tumour histology.

Immune checkpoint inhibitors are central to therapy for non-small cell lung cancer, but many patients do not respond^5^. Current predictive immunotherapy biomarkers include tumour PD-L1 expression^6^ and tumour mutation burden (TMB)^6^ but both provide limited prognostic information^5^.

Beyond mutation-driven cell proliferation, the tumour microenvironment (TME) is increasingly recognised to have profound effects on cancer outcomes as diverse cell types within a tumour may support cancer cell survival, spread, and escape from immunosurveillance^7^.

Despite histological features of LUAD being primary indicators of patient outcomes they are not typically included in the design of clinical trials. We therefore hypothesised that different histological patterns contain distinctive TMEs and immune features that are of importance for prognostic and therapeutic stratification. Here we characterize the histology of primary LUAD tumours through global gene expression and CIBERSORT immune profiling, in addition to mutation detection and SNP genotyping.

Global gene expression profiling detects biological activities of cells and tissues with immense power and accuracy. It can systematically detect underlying mechanisms and pathways in tumours, with the potential to target therapies. Gene expression is co-ordinated in molecular networks within cells and tissues^8^. Network analysis detects closely co-regulated modules of genes that, in contrast to single-cell sequencing, reveal integrated functions of tissue and cellular transcriptomes^9^. Individual networks can be associated with disease and related phenotypes through their module eigenvectors^10^, and their function inferred from their gene content. At the same time, immune cell types can be quantified in tumour samples by their distinctive transcriptional signatures^11^.

We have therefore uncovered gene co-expression modules with network analyses to understand molecular and immune features of LUAD morphology. We relate these modules of coregulated genes to standardised histologically defined features and identify immune cell content of the different growth patterns.

## Results

### Clinical and histologic characteristics

The study included 89 cases of LUAD, with tissue samples taken at surgery with curative intent. None of the patients had received chemotherapy or immunotherapy at the time of tumour resection. Four patients were discovered to have metastatic disease at the time of surgery and subsequently classified as stage IV TNM. The median age of patients at diagnosis was 68 years (range 45-89). Most tumour samples were acinar in pattern (37/89, 42%), with a lepidic pattern in 13 tumours (15%), papillary in 13 (15%), solid in 12 (13%) and micropapillary and cribriform in 7 each respectively (8%). Forty-one tumours (46%) were Grade 3 (G3), 35 (39%) G2 and 13 (15%) G1 (Table 1).

**Table 1.** Subject and tumour characteristics.

| Pattern | N | Grading | % Male (N) | Mean age (SD) | Never smoked (%) | Months survival by pattern (median/IQR) | Months survival by grade (median/IQR) |
| --- | --- | --- | --- | --- | --- | --- | --- |
| **Lipidic** | 13 | G1 (n=13) | 46.2% (6) | 69.9 (8.3) | 15% | 59 (27.5) | G1=59 (27.5) |
| **Papillary** | 13 | G2 (n=11), G3 (n=2) | 30.8% (4) | 68.3 (5.7) | 8% | 33 (51.5) | G2=56 (44.5) |
| **Acinar** | 37 | G2 (n=24), G3 (n=13) | 43.2% (16) | 67.3 (9.5) | 22% | 57 (41.5) | G3=50 (52) |
| **Cribriform** | 7 | G3 (n=7) | 42.9% (3) | 70 (13.9) | 0% | 62 (73) |  |
| **Micropapillary** | 7 | G3 (n=7) | 57.1% (4) | 66.6 (11.1) | 14% | 54 (24) |  |
| **Solid** | 12 | G3 (n=12) | 75% (9) | 68.4 (8.9) | 0% | 28 (53.3) |  |

The median survival of all subjects was 56 months (range 0-86, inter-quartile range (IQR) 46.5), with lepidic cases having the best survival (median 59 months, range 1-86, IQR 27.5) and solid the poorest (median 28 months, range 0-85, IQR 53.5) (Table 1). There was no significant difference in survival in this modest sample size, either by histological pattern or by tumour grade (Supplementary Figure 1A and 1B). Thirty (34%) of subjects were current smokers, 46 (52%) ex-smokers and 12 (13%) had never smoked. The smoking rates did not differ between histological patterns.

### Genetic abnormalities

#### Mutation frequencies

Seventy-nine of 83 sequenced tumours had detectable variants and paired normal tissue for germline analysis. Further filtering, to only include variants with Variant Allele Frequency (VAF)>5, led to subsequent mutation analyses being focused on 76 tumours. Figure 1A shows the oncoplot for the top 30 mutated genes with VAF>5, while Supplementary Table 2 shows the list of all detected variants (irrespective of VAF).

**Figure 1.**
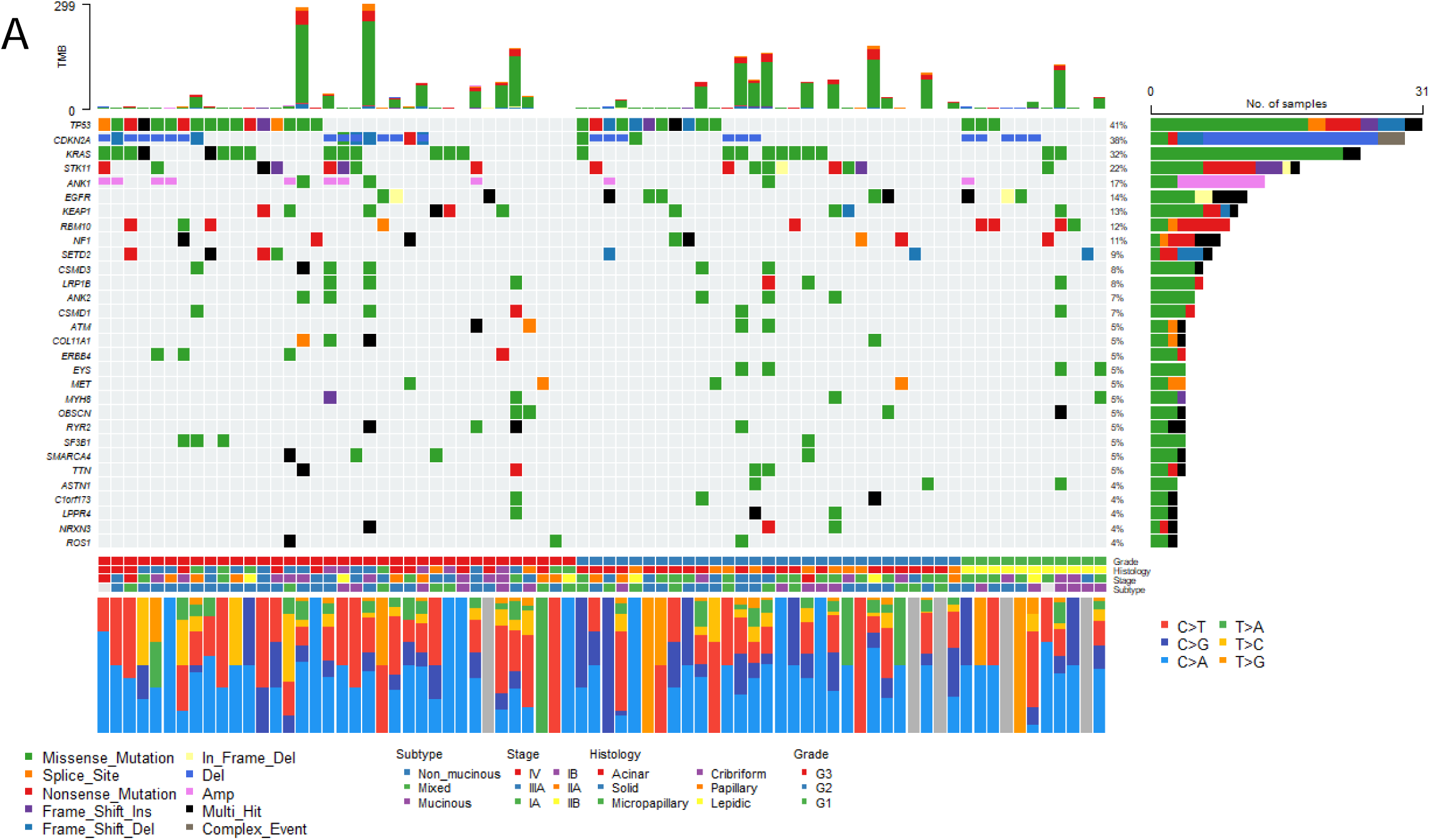
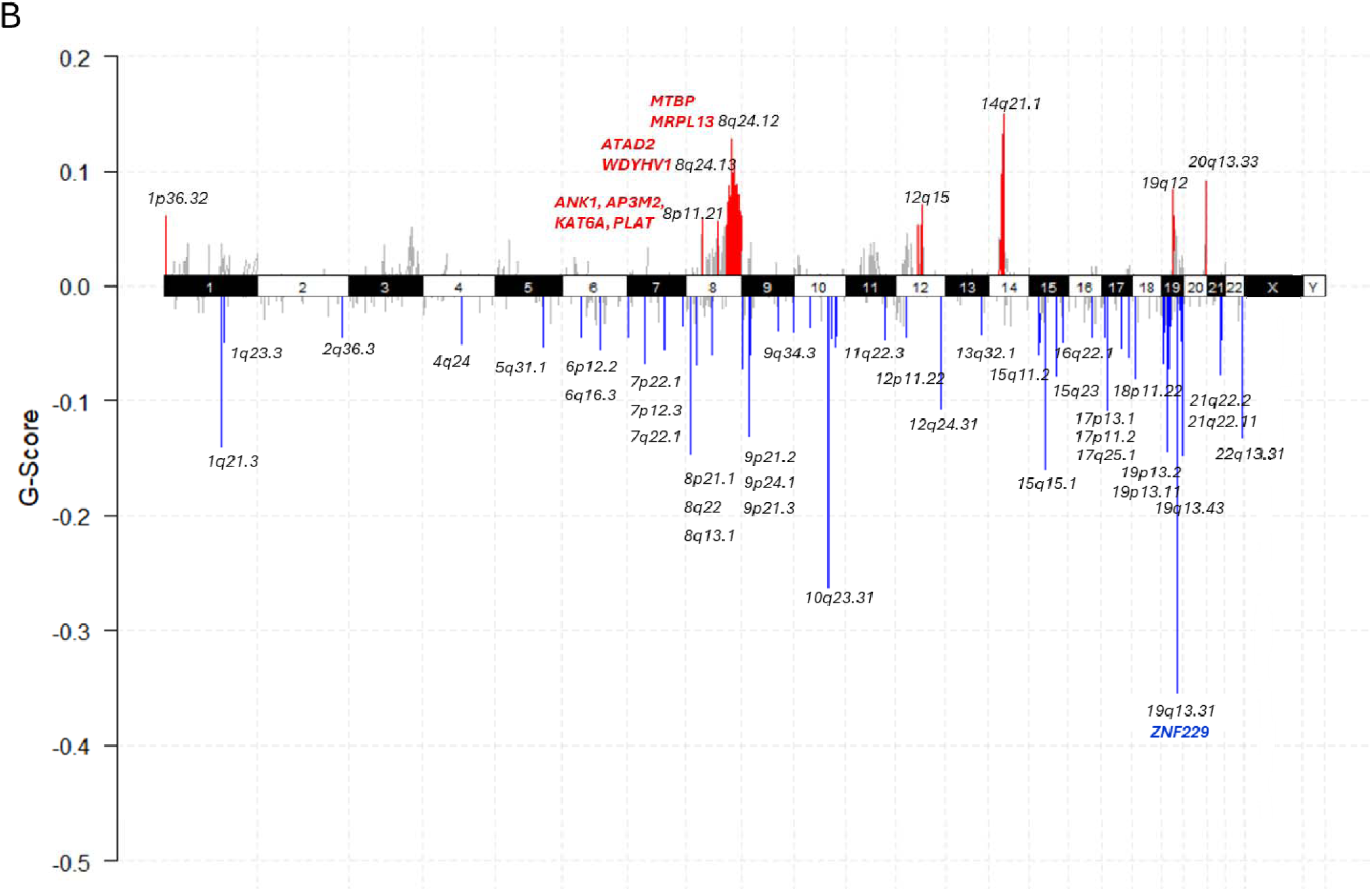
Mutations and copy number variation. A) Oncoplot of top 30 mutated and deleted or amplified genes. Patients are grouped according to the histologic grading (Grade1-Grade3). Upper panel shows 73 (who had VAF>5 out of 76) patients with mutations/ copy number alterations (mutation/copy number frequencies were calculated as percentages by dividing the number of patients with events to 76; a full list of variants is presented in Supplementary Table 2); middle panel shows classification of tumours based on histological grading, histological pattern, tumour stage and tumour subtype; bottom panel shows proportion of transition and transversion classes for each patient. B) Significant regions of amplification and deletion from GISTIC analysis of 79 subjects, showing genes in main peaks. Deletions are shown in blue and amplifications in red for peak regions that pass G-score (defined as the negative logarithm of the probability of observing a candidate copy-number segment with given amplitude and frequency, provided the background copy-number alteration rate) and q-bound (<0.05) threshold cut-offs.

Mutations affecting *TP53* were found in 41% of tumours and were present in 66.7% of the solid type and not detected in cribriform tumours (Supplementary Figure 1C). *CDKN2A* mutations or deletions were found in 38% of the patients; *KRAS* mutations in 32%; *STK11* in 22% and *EGFR* in 14%, each without frequency differences between histological groups (Supplementary Figure 1C).

*EGFR* mutations accompanied acinar (22.6%), lepidic (27.3%) and micropapillary patterns (9.1%). *EFGR* mutations did not distinguish tumour grade, with three mutations found in Grade 1 (G1) tumours, five in G2 and three in G3. Mutational exclusivity appeared between *EGFR* and *KRAS*, as expected (Supplementary Figure 1D)^12^.

Other potential actionable therapeutic targets with, included four MET mutations in G2 and G3 tumours (three out of four in the acinar pattern and one in papillary pattern); and one ERBB2 (Her2) in-frame insertions in a lepidic G2 tumour and one BRAFV600E mutation in a G3 tumour and a second one, with VAF<5 in a G2 tumour (Supplementary Table 2 and Supplementary Figure 1E).

*KRAS*, *TP53* and *CDKN2A* abnormalities were more common in ex-smokers and current smokers than in never smokers (Table 2). As expected, *EGFR* mutations were most prevalent in never-smokers (45.5%)^13^.

**Table 2.** Common mutations and smoking status.

| *Mutated Gene:* | *EGFR* |  | *KRAS* |  | *TP53* |  | *CDKN2A* |  |
| --- | --- | --- | --- | --- | --- | --- | --- | --- |
|  | N | % | N | % | N | % | N | % |
| **Never-smokers** | 5 | 45.5 | 1 | 4.2 | 3 | 9.7 | 3 | 10.3 |
| **Ex-smokers** | 4 | 36.4 | 14 | 58.3 | 15 | 48.4 | 15 | 51.7 |
| **Current smokers** | 2 | 18.2 | 8 | 33.3 | 13 | 41.9 | 10 | 34.5 |
| **Unknown** | 0 | 0.0 | 1 | 4.2 | 0 | 0.0 | 1 | 3.4 |
| **Total** | **11** | **100** | **24** | **100** | **31** | **100** | **29** | **100** |

#### Mutation spectra

Ninety-five % of mutations were single nucleotide polymorphisms (SNPs) producing missense, splice site or nonsense substitutions. Base substitutions were most often C>A transversions (48.6%) followed by C>T transitions (28.1%) (Supplementary Figure 2A). Current smokers showed an excess of C>A transversions over never smokers (OR= 1.9, *P*= 0.02, 95%CI [1.06-3.5]), with C>T transitions more common in never-smokers (OR= 1.8, *P*= 0.03, 95%CI [0.99-3.24]) (Supplementary Figure 2B). A mutational spectrum dominated by C>A mutations is consistent with exposure to the polycyclic aromatic hydrocarbons in tobacco smoke^14^, whereas C>T mutations probably follow oxidative DNA damage^15^.

#### Copy number aberrations and tumour mutational burden

Full details of the spectrum of copy number aberrations present in the LUAD tumours (shown in Figure 1B) is given in Supplementary Table 3. Deletion of segments was more common than their amplification (41 deleted vs 8 amplified, q-value <0.05). The most common deleted locus was 19q13.31 (gene *ZNF229*) found in 41/79 samples and the most frequently amplified loci were 8q24.12 (genes *MTBP* and *MRPL13*) and 8q24.13 (genes *ATAD2* and *WDYHV1*) detected in 28/79 samples each. None of these abnormalities correlated with tumour type or grade.

There was no association between TMB or copy number burden (CNB) with tumour grading in our subjects, although we observed a tendency towards increased TMB in poorly differentiated tumours (Grade 3) compared to well and moderately differentiated tumours (Grades 1 and 2) (Supplementary Tables 4 and 5).

Tumour mutation burden (TMB) is a possible indicator of neoantigen formation and predictor of response to immunotherapy^16^. Negative and positive correlations were respectively present between TMB and percentage of lepidic pattern (Spearman r=-0.28, *P*=0.01) and percentage of solid pattern (Spearman r=0.29, *P*=0.009), suggesting higher levels of antigenic peptides may accompany the high-risk solid pattern (Supplementary Figure 2C).

### Global gene expression with network analyses

After removal of transcripts with low abundance (below the median of all samples), the Affymetrix gene expression data was subjected to weighted gene correlation network analysis (WGCNA)^10^. Twelve modules of co-expressed genes were identified (by convention named with colours, Figure 2, Supplementary Table 6) and correlations sought between the network eigenvectors and the percent tumour type (Figure 2). These showed distinct modules of co-expression associated with each histological class. Active processes in individual modules could be inferred by inspection of hub genes (Supplementary Table 6). Likely cells of origin for network hubs were inferred from single-cell gene expression maps from lung tissues in the Human Protein Atlas^17,18^.

**Figure 2:**
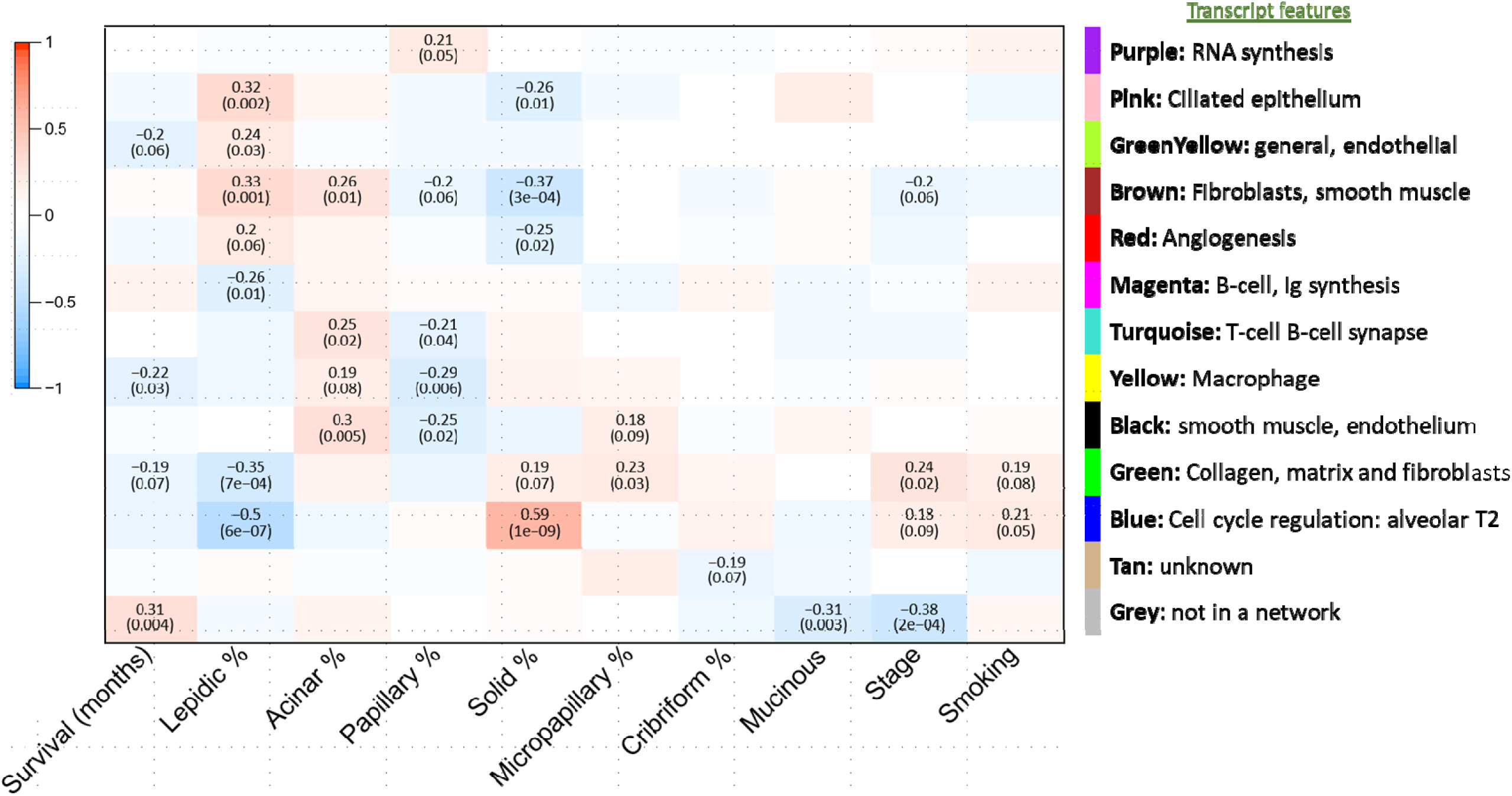
Network module content and associations. The matrix shows correlation between modules eigenvectors and histologic features of the tumours. Weighted gene correlation network analysis (WGCNA) identified and named (colours) twelve modules of co-expressed genes.

The results are summarised in Table 3 and detailed further below.

**Table 3.** Summary of associations between histology and immune and gene-expression features.

| Pattern | Proportion | Transcriptome module features | CIBERSORT Immune features |
| --- | --- | --- | --- |
| **Lepidic** | 14.6% | Ciliated epithelium, fibroblasts (brown module), angiogenesis, endothelium | Resting mast cells |
| **Papillary** | 14.6% | Small nuclear RNA synthesis | Activated mast cells |
| **Acinar** | 41.6% | Fibroblasts (brown), macrophages, smooth muscle, endothelium, T-cell B-Cell synapse | NK cells resting, M1 macrophages |
| **Cribriform** | 7.9% |  | CD8+ T-cells, resting mast cells |
| **Micro-papillary** | 7.9% | Smooth muscle, endothelium, collagen, matrix and fibroblasts (green module) | Activated dendritic cells |
| **Solid** | 13.5% | Alveolar, collagen, matrix and fibroblasts (green), cell cycle | Activated CD4+ memory, CD8+, M2 macrophages |

Associations with solid and lepidic cell content were generally opposing in directionality (Figure 2). The blue module was most strongly associated with solid histology (r=0.59, *P*=1E-09), and its hub genes included *TPX2*, *MYBL2* and *PLK1* (module membership MM=0.94 to 0.92, *P*=1.2E-42 to 1.2E-37) (Supplementary Table 6_Blue module (solid associated)), each of which have central roles in cell cycle progression and poor outcomes in numerous cancers. Inspection of hub gene single-cell expression from lung tissues in the Human Protein Atlas^17,18^ was consistent with an alveolar origin for solid lung cancers.

Lepidic pattern by contrast strongly associated with the pink module (r=0.32, *P*=0.002), which contained numerous genes encoding the components of cilia, such as *DNAI1*, *CFAP43*, *CFAP57*, *ZBBX*, *TEKT1*, *DNAH12*, *DNAAF1*, *CFAP61* (MM=0.96 to 0.93, *P*=2.9E-49 to 2.1E-39) (Supplementary Table 6_Pink (lepidic and mucinous)). Also significantly associated with lipid features was the brown module (r=0.33, *P*=0.001) with hubs active in the tissue matrix (*ITGA8*, *A2M*, *CPED1*) and in angiogenesis (*ANGPT1*, *SLIT3*, *SLIT2*) (MM ranging from 0.9 to 0.84, *P=*2.8E-33 to 5.2E-25); whilst the red module showed a trend towards positive association (r=0.2, *P*=0.06) with hubs for angiogenesis including *CDH5*, *TEK*, *MYCT1* and *CLEC14A* (MM=0.93 to 0.86, *P*=1.2E-38 to 1.3E-27) (Supplementary Table 6_Red (lepidic)).

An acinar pattern was also positively associated with the tissue matrix brown module (r=0.26, *P*=0.01, Figure 2). Concomitant correlation of acinar histology with the black module (r=0.3 *P*=0.005) indicated other fibroblast and smooth muscle elements (Supplementary Table 6_Black (acinar)).

Co-ordinated immune processes were found in the turquoise and magenta modules. The acinar-associated turquoise module (r=0.25, *P*=0.02) contained multiple elements of the T-cell B-cell synapse including *PTPRC, BTK, CD37, CD38, ZAP70, CD3D, CD3G*, and *CD2* (MM=0.94-0.85, *P*=9.8E-43 to 7.4E-26) (Supplementary Table 6_Turquoise (T-cell receptor and B cells). The magenta network encoded B-cell development (e.g. *FCRL5, LAX1, MZB1*) and immunoglobulin synthesis, containing multiple IGH, kappa and lambda variable regions (MM=0.96 to 0.75, *P*=1.7E-47 to 1.5E-17) (Supplementary Table 6_Magenta (B cell)), and was negatively associated with lepidic tumours (Figure 2).

Papillary tumours were associated with the purple module, with hub genes containing multiple small nucleolar RNAs (Supplementary Table 6_Purple (papillary)), which are intron encoded and enact posttranscriptional maturation of ribosomal and other cellular RNAs^19^.

### Immune cell content

We quantified immune cell subsets within tumour transcriptomes using CIBERSORT^11^, which applies vector regression with prior knowledge of expression profiles from leukocyte subsets to estimate tumour immune composition. Plasma cells, memory CD4 resting T-cells, resting mast cells and M2 macrophages were amongst the most abundant cells detected (Supplementary Figure 3A).

The distribution of different cells varied greatly between normal and tumour samples (Supplementary Figure 3B) and between different histological patterns (Supplementary Figure 3C) and with some mutation and structural aberrations (Supplementary Table 7). Principal component analyses (PCA) identified numerous clusters of these variables that related strongly to variance within the data (Supplementary Table 8).

Formal modelling with regression analysis identified highly significant independent associations with different histological types (Table 4, Supplementary Table 9). Solid histology was predicted by the abundance of CD4 memory activated T-cells (R^2^=15.4%, *P*=0.000), CD8 T-cells (additional R^2^=6.8%, *P*=0.007), mutations in *TP53* (additional R^2^=4.0%, *P*=0.034), and the abundance of M2 Macrophages (additional R^2^=3.4%, *P*=0.047). By contrast lepidic tumours were associated with the abundance of resting mast cells (R^2^=10.7%, *P*=0.002).

**Table 4.**
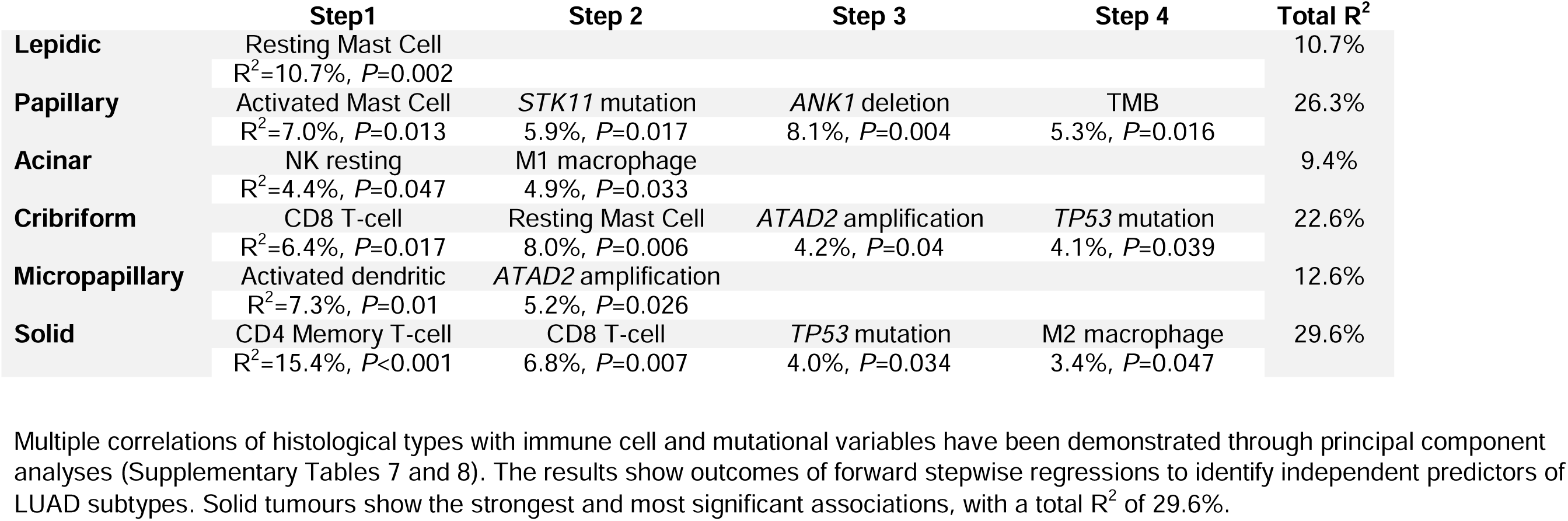
Immune cell and mutation predictors of LUAD histological classifications. Multiple correlations of histological types with immune cell and mutational variables have been demonstrated through principal component analyses (Supplementary Tables 7 and 8). The results show outcomes of forward stepwise regressions to identify independent predictors of LUAD subtypes. Solid tumours show the strongest and most significant associations, with a total R^2^ of 29.6%.

|  | Step1 | Step 2 | Step 3 | Step 4 | Total R<sup>2</sup> |
| --- | --- | --- | --- | --- | --- |
| **Lepidic** | Resting Mast Cell<br>R<sup>2</sup>=10.7%, P=0.002 |  |  |  | 10.7% |
| **Papillary** | Activated Mast Cell<br>R<sup>2</sup>=7.0%, P=0.013 | STK11 mutation<br>5.9%, P=0.017 | ANK1 deletion<br>8.1%, P=0.004 | TMB<br>5.3%, P=0.016 | 26.3% |
| **Acinar** | NK resting<br>R<sup>2</sup>=4.4%, P=0.047 | M1 macrophage<br>4.9%, P=0.033 |  |  | 9.4% |
| **Cribriform** | CD8 T-cell<br>R<sup>2</sup>=6.4%, P=0.017 | Resting Mast Cell<br>8.0%, P=0.006 | ATAD2 amplification<br>4.2%, P=0.04 | TP53 mutation<br>4.1%, P=0.039 | 22.6% |
| **Micropapillary** | Activated dendritic<br>R<sup>2</sup>=7.3%, P=0.01 | ATAD2 amplification<br>5.2%, P=0.026 |  |  | 12.6% |
| **Solid** | CD4 Memory T-cell<br>R<sup>2</sup>=15.4%, P<0.001 | CD8 T-cell<br>6.8%, P=0.007 | TP53 mutation<br>4.0%, P=0.034 | M2 macrophage<br>3.4%, P=0.047 | 29.6% |

Acinar pattern was associated with resting NK cells (R^2^=4.4%, *P*=0.047) and M1 Macrophages (additional R^2^=4.9%, *P*=0.033). Papillary pattern was associated with activated mast cells (R^2^=7.0%, *P*=0.013) and deletion of *ANK1* (additional R^2^=8.1%, *P*=0.004). Cribriform patterns were associated with resting CD8 T-cells (R^2^=6.4%, *P*=0.017) and resting mast cells (additional R^2^=8.0%, *P*=0.006). Micropapillary patterns were associated with activated dendritic cells (R^2^=7.0%, *P*=0.013).

### Survival-associated transcripts

Patient survival was negatively associated with the yellow module (r=-0.22, *P*=0.03), which contained macrophage-associated hub-genes *FPR3*, *CD68*, *HCK*, *SIRPA* and *SPI1*. (Supplementary Table 6_Yellow module (survival)). This confirms the known importance of macrophages in progressive and aggressive cancer^20^.

Far stronger negative associations, however, were found between individual transcripts and patient survival (Table 5). These transcripts belonged to the green fibroblast or to the yellow macrophage networks (Supplementary Table 6_Green (micropapillary) and Yellow module (survival)), and included recognised therapeutic or prognostic targets including *CTSL1*^21^ (Hazard Ratio (HR)=2.72, *P<*0.001), *KRT17 (*HR=1.43, *P<*0.001), *ADORA3*^22^ (HR=1.81, *P=*0.005), *EPB41L3*^23^(HR=2.36, *P=*0.004), and *IL2RA*^24^ *(HR=1.9*, *P=*0.001). Multivariate analyses revealed independent effects for *KRT17*, *TNS4*, *FUT2*, *SERPINE1*, *PTGER2*, *PLIN2* and *H19* (Table 5 – genes with asterisks and Supplementary Table 6_Survival top hits), suggesting combined prognostic value from these transcripts. Their Kaplan-Meier curves are shown in Figure 3.

**Table 5.**
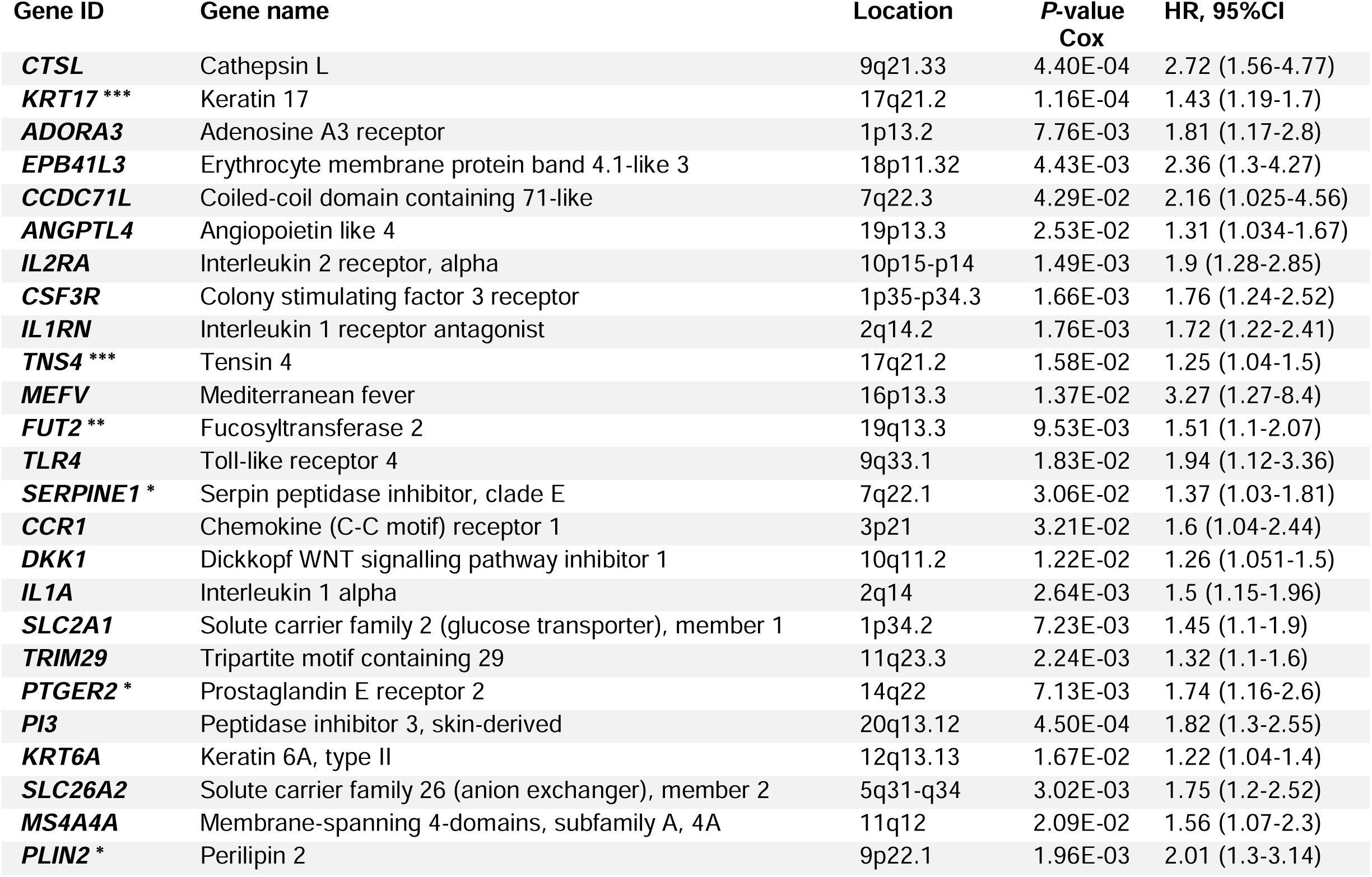
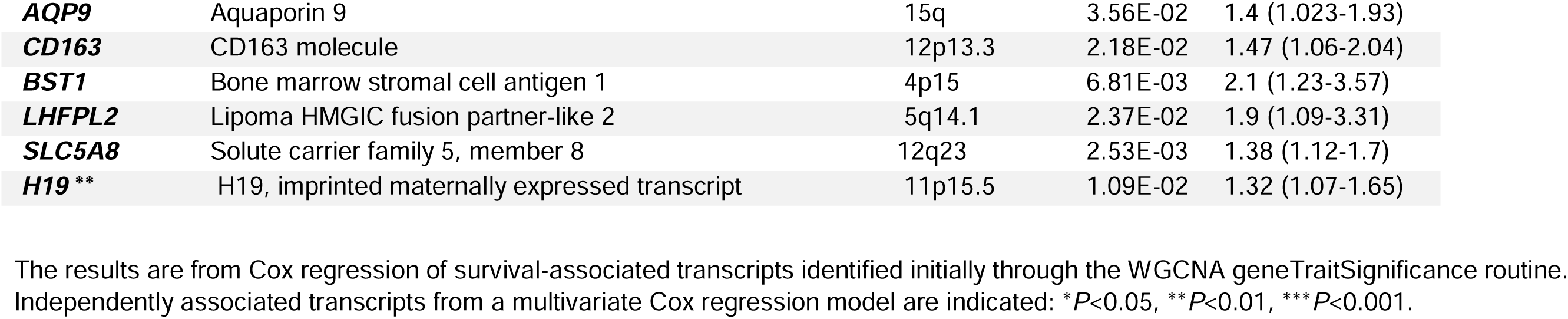
Survival associated transcripts. The results are from Cox regression of survival-associated transcripts identified initially through the WGCNA geneTraitSignificance routine. Independently associated transcripts from a multivariate Cox regression model are indicated: \**P*<0.05, \*\**P*<0.01, \*\*\**P*<0.001.

**Figure 3:**
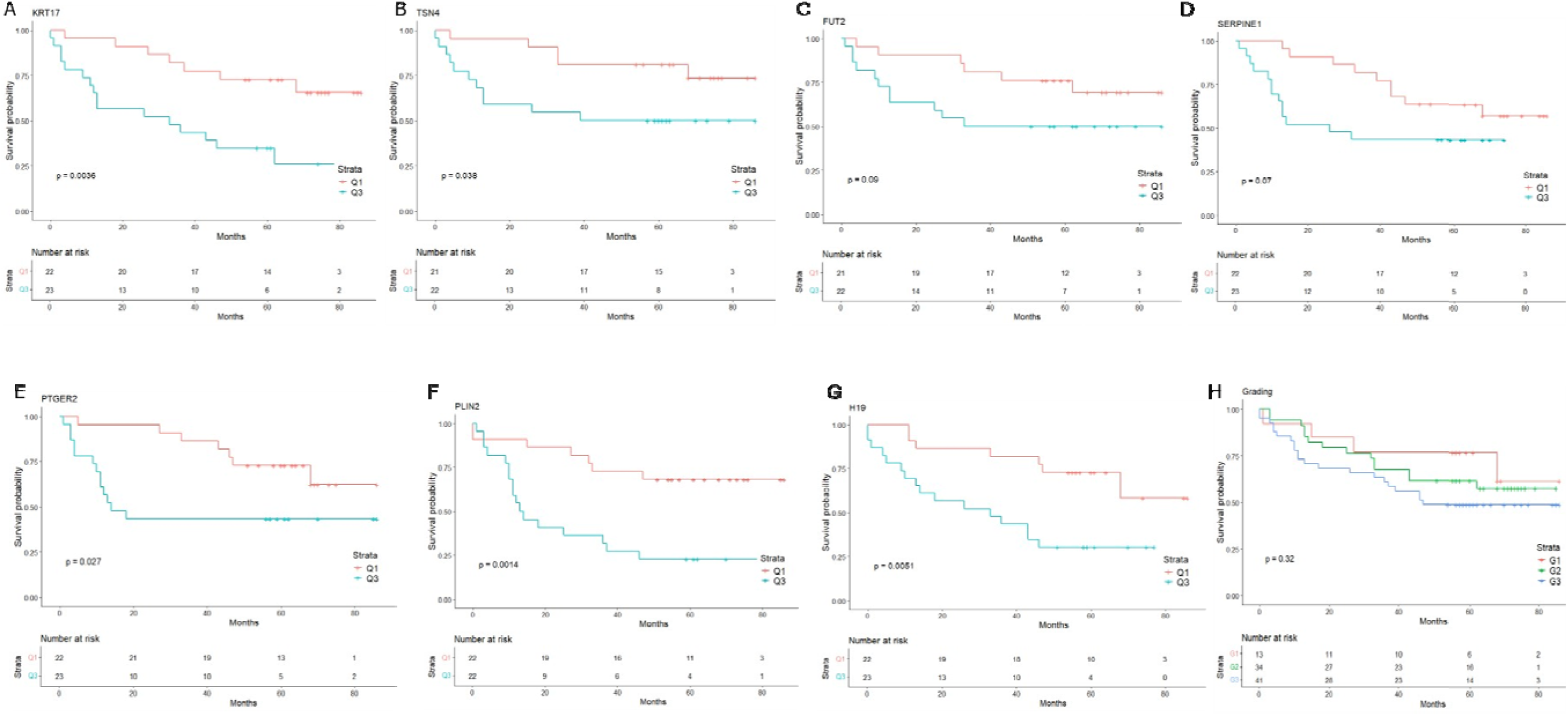
Kaplan-Meier curves for transcripts independently associated with survival. Kaplan-Meier curves for transcripts that maintained statistical significance with survival after multi-variate Cox regression. Survival curves compare the 25% and 75% quartile of transcript abundance for each transcript: A) *KRT17*, B) *TSN2*, C) *FUT2*, D) *SERPINE1*, E) *PTGER2*, F) *PLIN2* and G) *H19.* H) Survival according to histological grade shown for comparison.

*CD274* (PD-L1) is the principal target for immunotherapy, and its gene was found in the yellow macrophage module (MM=0.69, *P*=4.7E-14 Supplementary Table 6_Yellow module (survival)), but its expression was not associated with survival (r<0.001, *P*=0.99). The gene for PD1 (*PDCD1*) itself was below the threshold for detection in the transcriptomic data.

## Discussion

Our study shows that WHO-defined LUAD histological patterns contain deep levels of molecular complexity that entrain different cells of origin, tumour microenvironments and immune cell content. These elements are not related simply to mutations or structural genomic abnormalities.

### Tumour-specific gene expression and immune cell profiles

Solid and lepidic predominant tumours are at different ends of the LUAD grading spectrum^25,26^, and in our data we identified associations with co-expression modules that were also opposing in direction.

Solid pattern tumours were strongly related to the blue module, the hub genes of which are consistent with an alveolar cell origin. Solid histology was strongly and independently predicted by the abundance of CD4 memory activated T-cells (R^2^=15.4%), and CD8 T-cells (additional R^2^=6.8%). CD8^+^ T-cells are considered the major element of the immune response to tumours^27^, and CD4^+^ T-Cells may direct sustained immune reactions^28^.

Our findings agree with the previous observation of adaptive immune resistance, higher cytotoxic activity, and enhanced immunogenicity within solid-predominant histology^29^, and our findings support the suggestion that solid tumours may selectively benefit from immunotherapy with check-point blockade^29^.

Anti-PD1 or anti-PD-L1 antibodies are currently used to stratify LUAD patients for checkpoint immunotherapy. However, in our data PD1 transcripts were below the threshold for detection for our arrays, and PD-L1 (*CD274*) was a member of the yellow macrophage model rather than associated with any tumour subtype. This concurs with the general observation that the predictive value of PD-L1 expression in clinical trials is suboptimal^30^. Tumour mutational burden (TMB) also may predict susceptibility to immunotherapy^31^, but we did not find any associations between TMB and immune profiles in our data.

Ciliary origins for lepidic tumours were strongly suggested by the hub genes of their associated pink module, although previously it has been assumed that LUAD in general arises from alveolar cells^32^. Lepidic and acinar histology were both positively associated with the brown fibroblast element network, and additional smooth muscle and endothelial elements in acinar tumours are indicated by a black module association.

Fibroblasts are a central component of the TME in many cancers, exhibiting profound influences on progression through extensive interactions with cancer cells and other stromal elements^7^. Therapies directed against fibroblasts are however problematic, attributed in part to extensive heterogeneity amongst fibroblast subsets within different tumours^7^. Distinct differences in associations with tissue matrix modules suggest that histological classification may help stratify trials of anti-fibroblast therapy. They also identify network hubs in the brown (*CACNA1C, PRKG1, BNC2*) and green (*COL5A2, ADAMTS12, SULF2, THBS2*) modules that are potential biomarkers for fibroblast subsets.

Angiogenesis is a another complex process that plays a crucial role in tumour growth, invasion, and metastasis^33^. The benefits of anti-angiogenic therapies are also difficult to predict^33^. In our results, solid tumours were negatively associated with angiogenic genes in the red module, suggesting a mechanism for therapy resistance in this subtype.

Papillary tumours were uniquely related to high expression of small nucleolar RNAs in the purple module. snoRNAs are intron encoded and enact post-transcriptional maturation of ribosomal and other cellular RNAs^19^. Mutations and aberrant expression of snoRNAs have been reported in cell transformation, tumorigenesis, and metastasis^19^, and RNA splicing aberration can generate actionable neoantigens. Circulating snoRNAs are potential biomarkers for LUAD progression^34^, and may be of prognostic benefit in papillary tumours.

Distinctive immune cell contents were enriched in each histological class, for example NK cells and M1 macrophages in acinar LUAD; activated mast cells in papillary tumours; activated dendritic cells in micropapillary LUAD; and resting CD8 T-cells and resting mast cells in cribriform tumours. Our results confirm an earlier study of 174 resected stage I-III LUAD tumours classified by histologic pattern and level of differentiation. Genomic analysis revealed high TMB in poorly differentiated tumours, but no association with oncogenic drivers. Immune analyses showed infiltration by cytotoxic CD8 T-cells, activated CD4 T-cells, and pro-inflammatory (M1) macrophages^35^, in line with our findings.

The role that these individual immune associations may play in subtype tumour progression are not yet known, but they strongly suggest histological stratification will be informative in trials of immunotherapy.

It is of interest that two modules of coordinated immune processes (multiple elements of the T-cell B-cell synapse (including CD8A) in the turquoise module and B-cell-immunoglobulin synthesis in the magenta module) are resident in the matrix of normal airway mucosa^36^. In the current study, T-cell B-cell interactions were positively associated with acinar tumours, consistent with a response to neoantigens. The negative correlation with the immunoglobulin synthesis module in lepidic tumours might suggest low antigenicity.

### Mutations

The frequency of individual mutations in our cases was typical for LUAD, with *TP53* mutation most common followed by *CDKN2A* mutations and deletion, *KRAS* mutations, *STK11* and *EGFR* mutations. Lesions in individual genes did not predict histology, although *TP53* mutations were more frequent in G2 and G3 tumours, while *EGFR* was infrequent in cribriform, micropapillary and solid pattern tumours. As expected, *KRAS*, *TP53* and *CDKN2A* abnormalities were more common in ex-smokers and current smokers and *EGFR* mutations prevalent in the never smokers^13^. Mutational exclusivity appeared between *EGFR* and *KRAS,* which has previously been attributed to synthetic lethality^37^. Mutations in individual genes did not significantly predict patient survival in our data.

A previous study of mutation profiles in 604 patients with surgically resected LUAD^3^ has shown TMB to be elevated in high-grade tumours, and this was subsequently confirmed^35^. That we did not reproduce the reported associations with TMB with recurrence is attributable to our smaller sample size, but our results illustrate the limited effect size of histological/mutational relationships.

### Survival

Histological patterns were also only weakly associated with survival, partly attributable to the limited number of subjects. We recognise that we classified histological classes with a focus on the small portion of the tumour submitted to genomic analyses, and we expect that the predominant pattern in the specimen overall would give better prognostication.

Our data suggest specific markers that may predict outcomes or guide therapy. We found the strongest relationships to survival were with elevated expression of inflammatory genes. Strikingly, many of these are potential therapeutic or prognostic targets, including *CTSL1*^21^, *KRT17*^38^, *ADORA3*^22^, *EPB41L3*^23^, *CCDC71L*^39^ and *IL2RA*^24^. Replication in the online web interface kmplot showed a significant correlation between longer survival and low gene expression, except for *EPB41L3* and *CCDC71L* where the significant correlation with survival was seen with high gene expression.

In general, the transcripts listed above tended to belong to the yellow (macrophage) and green (fibroblast) modules. Inflammation is recognised to drive tumour initiation, growth, progression, and metastasis, and macrophages stimulate angiogenesis, tumour cell invasion, and intravasation at the primary site^20^. The significant content of M2 Macrophages in solid tumours may indicate their suitability for targeted anti-inflammatory therapies.

## Conclusions

Overall, our results confirm marked differences in cells of origin, matrix activities and immune cell content across the LUAD spectrum. The results suggest multiple diagnostic markers that may augment the WHO histology classification. Even before full understanding of the underlying processes, it is reasonable to consider whether stratification according to WHO histology would add to the precision of clinical trials.

It is also possible that some proteins may help in automated histology or in circulating tumour cell cytology. These include *TPX2*, *MYBL2* and *PLK1* for solid content, and cillial genes such as dyneins (*DNAI1*, *DNAH12*, *DNAAF1)* and cilia and flagella-associated proteins (*CFAP43*, *CFAP57*, *CFAP61*) for lepidic tumours. If genes we found to be associated with poor survival (*CTSL1*, *KRT17*, *ADORA3*, *EPB41L3*, *CCDC71L* and *IL2RA*) are confirmed, they could provide prognostic information that is independent of histological class.

Finally, a natural extension of these findings is to expand histological and genomic analyses with machine learning technologies that can detect far more than the human eye^40^. Our results also suggest the potential for spatial transcriptomic profiling to inform tissue context and cellular interactions^41^ in the clinical translation of advanced image analyses.

## Material and methods

### Subjects

Tumour and paired non-tumoral tissues were part of a previously described collection of patients with surgically treated non-small cell lung cancer, analysed for Y chromosomal deletions^42^. In the present study we selected 89 patients with LUAD that had Affymetrix microarray data, out of which 83 had matched whole exome or targeted capture sequencing data and 82 had SNP genotyping data. The study was approved by the Royal Brompton and Harefield Research Ethics Committee (RBH) NIHR BRU Advanced Lung Disease Biobank (NRES reference 10/H0504/9) and Brompton and Harefield NHS Trust Diagnostic Tissue Bank (NRES reference 10/H0504/29) [Discovery], and the Royal Brompton and Harefield Ethics Committee (REC reference number LREC 02-261) [Replication]. Never smokers were patients that smoked less than 100 cigarettes during their lifetime, ex-smokers were patients who quit smoking more than 6 months prior to surgery while current smokers were patients that still smoked or quit less than 6 months ago.

### Histology

Histological patterns were assessed by an expert pulmonary pathologist (AGN), and sampled areas of tumour were classed as lepidic, acinar, papillary, solid, micropapillary or cribriform. The highest percentage pattern was considered the predominant histologic subtype. Tumours were graded as described^1^ into well-differentiated (lepidic predominant with no or less than 20% high-grade patterns), moderately differentiated (acinar or papillary predominant tumours with no or less than 20% high-grade patterns) and poorly differentiated (any tumour with 20% or more of high-grade patterns). Tumours were additionally classified as non-mucinous, mucinous or mixed. To quantify associations, tumours were also classified by percentage of histological pattern. In general, ordinal class and pattern percentage as outcomes gave very similar results.

### Genomic analysis

#### Whole-exome sequencing and targeted capture sequencing

We performed whole-exome sequencing in 24 samples, and for the remaining 59 samples used a hybridisation capture panel that covered the entire coding region of fifty-two genes (Supplementary Table 1) encompassing 266,937kb. Details on analysis, data processing and selection of pathogenic somatic mutations have been previously published^43,44^. Briefly, sequencing libraries were prepared from DNA extracted from tumours and normal tissue samples. Libraries for whole exome sequencing were prepared using SureSelect XT Human All Exon V4 (Agilent, Santa Clara, USA) and 100bp paired end sequenced on a HiSeq2000 sequencer (Illumina, San Diego, USA). Targeted capture sequencing libraries were prepared using the SureSelect QXT Target Enrichment System (Agilent, Santa Clara, USA) according to the manufacturer’s protocols. Sequencing was performed on NextSeq500/550 platform (Illumina, San Diego, USA). WES FastQ files were trimmed and aligned against the Human Genome December 2013 assembly (GRCh37/hg19) with BWA mem. SAM files were sorted with GATK (version 3.7) and read alignments were removed with Picard (version 2.9.0) to obtain BAM files. Somatic mutations and InDels were identified with MuTect (version 1.16) and Scalpel (version 0.4.1) software. Functional effect was predicted with SnpEff (version 4.3), and addition of metadata was done with Genome MINIng (GEMINI, version 0.14-0.20) software. Variants were filtered by known impact with CADD scores. Only variants with CADD scores>=15 were selected. For downstream analysis, variants with VAF<5 were filtered out.

Somatic variants from WES and targeted capture sequencing were merged using command line and RStudio and used to generate Multiple Alignment Format (MAF) files as input for Maftools^45^.

#### SNP genotyping, copy-number analysis and copy-number burden

Eighty-two paired (tumour and normal) DNA samples were interrogated at Eurofins against Human Infinium Omni-Express-Exome v 1.6 Bead Chips (Illumina) arrays containing 958,497 SNP markers, using microarray technology. Seventy-nine tumours presented significant peaks after analysis. SNP genotyping was analysed through SNP clustering GenomeStudio software (ver. 2.0.4) to generate raw copy number data (LRR and BAF). ASCAT version 2.5.1 was used for GC correction and generated Log R ratios (LRR) that were processed using DNA copy version 1.56.0. Germline subtracted copy number segments were processed with GISTIC version 2.0.23 to call for significant CNA in somatic tumour segments. Copy number burden (CNB) was calculated from the segment size of amplifications and deletions per autosome size and defined as percentage of genome carrying copy number aberration. Tumour purity was ensured by histological examination.

#### Tumour mutation burden (TMB)

Tumour mutation burden for each malignancy was defined as the total number of somatic coding, base substitution and InDel mutations detected per Mega base (Mb) of genome examined. CallableLoci from GATK (version 3.7) was used to obtain callable bases applying the following criteria: a minimum read depth of 10 before a locus was considered callable, a minimum base quality of 20 (based on Phred scores) and a minimum mapping of 30 reads to count towards depth. When testing associations with TMB, potential effects of different sequencing methods were controlled for by regression analyses that included “method” as an independent factor.

### Affymetrix gene expression

Eighty-nine paired tumour samples were part of a previously described dataset^42^, selected here to have overlapping sequencing, SNP genotyping and gene expression data. CEL files were analysed with *limma* and RMA normalised to generate log_2_ gene expression data. Only named transcripts were included in downstream analyses.

### CIBERSORT

We derived the abundance for 22 types of immune cells from bulk tumour transcripts with CIBERSORT^11^, utilising a cell-specific signature matrix of 547 genes.

### WGCNA

A consensus network analysis of tumour and normal lung expression data was performed using step-by-step unsigned weighted gene correlation network analysis (WGCNA (version 1.51))^10^ employing a soft-thresholding power of 4, and scaling topological overlap matrices (TOM) for purposes of comparability (scaling parameter 0.95).

Adaptive branch pruning was performed using dynamicTreeCut (version 1.63-1), applying a minimum cluster size of 30, a maximum joining height of 0.995 and a deep split parameter of 2 (specifying the sensitivity to cluster splitting). Modules classified as too close in terms of the correlation of their module eigengenes were merged (maximum dissimilarity that qualifies modules for merging 0.25). Consensus modules were related to phenotypic traits through two-sided bi-weight mid-correlation (robustY=FALSE, maxPOutliers=0.05 as per recommended best practice for settings that include binary or ordinal variables) and compared with modules identified in tumour or unaffected tissue alone as calculated using equivalent computational parameters.

### Survival

Survival was measured in months between the date of operation and the date of death or last follow-up. Simple associations to transcript abundance were identified with geneTraitSignificance in WGCNA and then formally assessed with Cox proportional hazard regression with the coxph function from survival package (version 3.8-3) and through the online web interface http://kmplot.com/^46^. Univariate and multivariate analysis were carried out with cut-off auto-selection. Kaplan-Meier survival curves comparing the lower quartile (Q1) abundance with the upper (Q3) were run with the survfit function and plotted with ggsurvplot from the survminer package (version 0.5.1).

### Statistical analysis

Clinical factors were summarized as numbers, frequencies (percentages) or medians (with range and interquartile range (IQR)). Analyses were run in R (version 4.2.1) and SPSS (version 26). Significance tests were two-sided, and *P* < 0.05 was considered statistically significant.

## Data Availability

All data produced in the present study are available upon reasonable request to the authors

## Acknowledgements

We thank the patients for donating their tissues to our study. This study was supported by a joint Wellcome Senior Investigator Award to WOCC and MFM (WT096964MA and WT097117MA) and by the Asmarley Trust. Sample collection was supported by the NIHR Respiratory Disease Biomedical Research Unit at the Royal Brompton and Harefield NHS Foundation Trust. The views expressed in this publication are those of the authors and not necessarily those of the NHS, the National Institute for Health Research or the Department of Health. The funding sources had no involvement in study design, collection, analysis or interpretation of the data, or in submission for publication.

## Statement of author contributions

AN, CDS, and AM carried out mutation and copy number variant analyses, designed by AN, MFM and DMR. AN and MO conceived and carried out immune cell profiling from transcriptomic data. ES supervised collection of LUAD specimens and associated metadata, recorded histological classifications with AGN, extracted nucleic acids, and carried out Affymetrix microarray assays. EL planned and implemented tissue access from surgical specimens. SAGWO and LL led bioinformatic analyses of gene expression with input from WOCC, who also modelled histology associations. The study was conceived, designed and led by MFM and WOCC. All authors were involved in writing the paper and had final approval of the submitted and published versions.

## List of supplementary material online Information

Supplementary Figure 1

A) Kaplan-Meier survival curves based on histological pattern; B) Kaplan-Meier survival curves based on histological grading; C) Proportion of main LUAD genes in the whole cohort stratified on histological subtype; D) Somatic interaction in top 10 mutated genes; E) Oncoplot of main LUAD genes (VAF>5) grouped on histological subtype.

Supplementary Figure 2

A) MAF summary for the mutated genes (variants with VAF>5); B) Transversions and transitions analysis based on smoking status; C) Spearman correlations between LUAD histology, age, CNB and TMB.

Supplementary Figure 3

A) Distribution of immune cells in each tumour sample; B) Immune cell abundances in normal versus tumour tissue all samples (Wilcoxon tests of significance FDR corrected); C) Proportion of immune cells in each tumour histological subtype (Wilcoxon test, FDR corrected).

Supplementary Table 1: List of 52 lung cancer associated genes in the targeted sequencing panel

The targeted gene capture panel used for screening for LUAD mutations. The hybridisation capture panel included the entire coding region of fifty-two genes, containing 12,129 probes with a total coverage of 266,937kb.

Supplementary Table 2: List of all variants detected

Supplementary Table 3: List of significant copy number variants (CNVs)

Supplementary Table 4: Tumour mutation burden (TMB) and copy mutation burden (CNB)

Supplementary Table 5: Summary of TMB and CNB distributions in risk groups and patterns

Supplementary Table 6. Summary of hub transcripts in modules identified by weighted gene correlation network analysis (WGCNA)

Transcript names are given at the left, with brief functional descriptors for the most significant genes. Modules are listed at the top. The MM statistic for each module describes strength of association between transcript abundance and the module eigenvector (1=complete, 0=none) and p.MM the P-value for the association. Sorting the spreadsheet by MM values allows inspection of hub genes for each module.

Supplementary Table 7. Correlation matrix between histological type, genomic abnormalities, and derived immune cell content.

Supplementary Table 8. Principal component analysis for histological type, genomic abnormalities, and derived immune cell content.

A) Tumours classified by % histology; B) Tumours classified as histology class.

Supplementary Table 9. Logistic regression analyses of mutation spectrum, immune cell content and histological class.

