## Supplementary Figures for "Lung adenocarcinoma WHO histological classes contain distinct immune cell profiles"

### Supplementary Figure 1

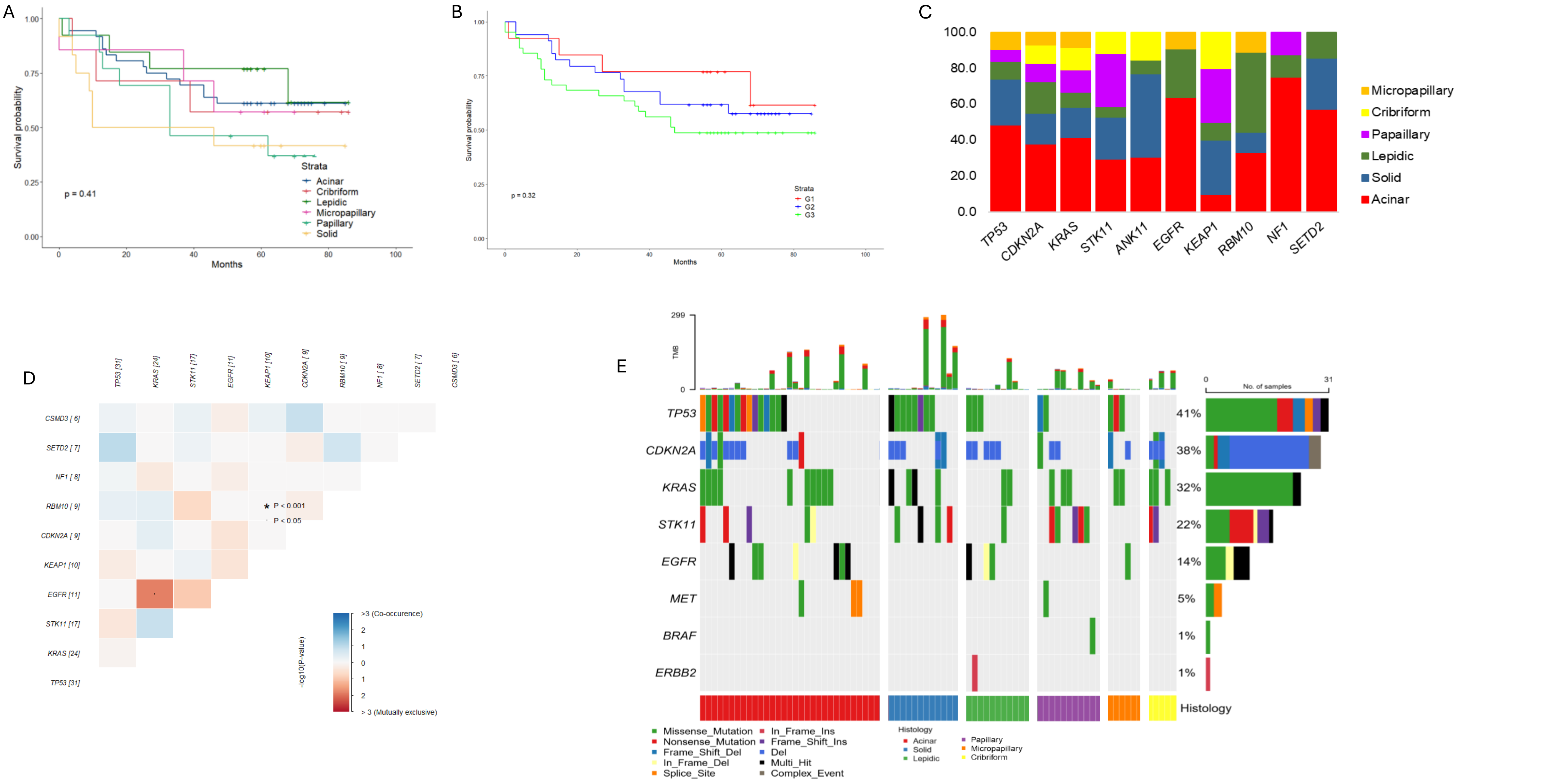


A) Kaplan-Meier survival curves based on histological pattern; B) Kaplan-Meier survival curves based on histological grading; C) Proportion of main LUAD genes in the whole cohort stratified on histological subtype ; D) Somatic interaction in top 10 mutated genes; E) Oncoplot of main LUAD genes grouped on histological subtype (VAF>5).

Supplementary Figure 2

A

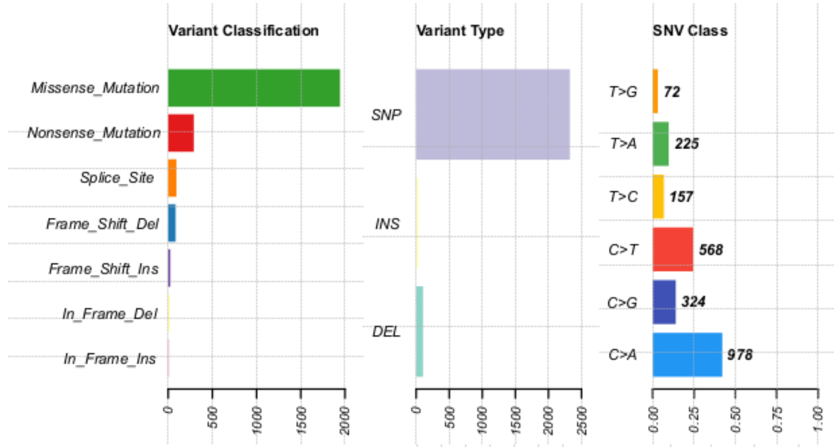


B

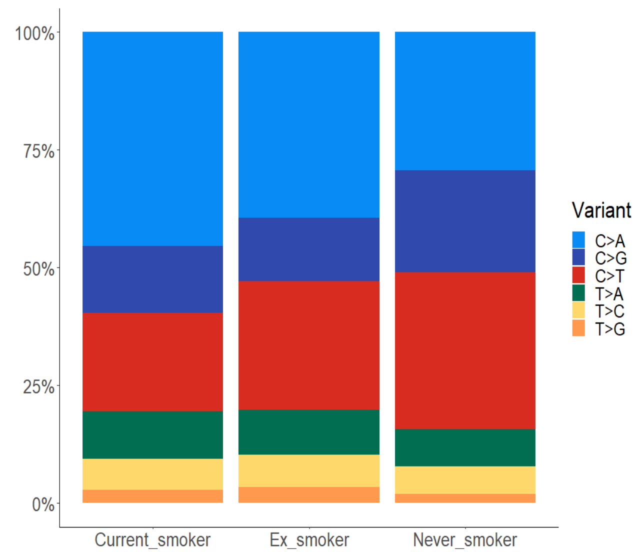


C

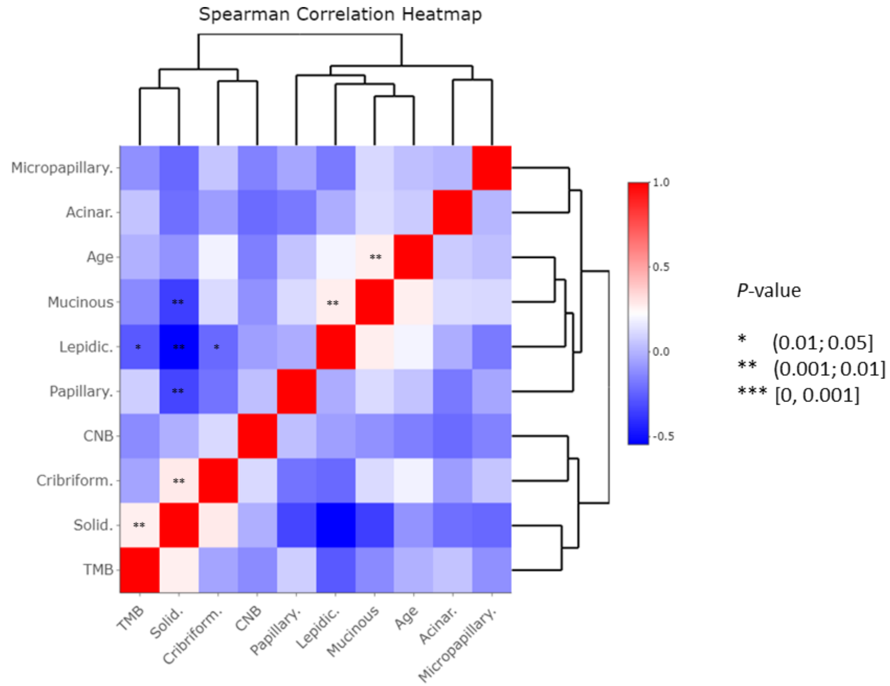


A) MAF summary for the mutated genes (variants with VAF>5); B) Transversions and transitions analysis based on smoking status; C) Spearman correlations between LUAD histology, age, CNB and TMB.

### Supplementary Figure 3

A

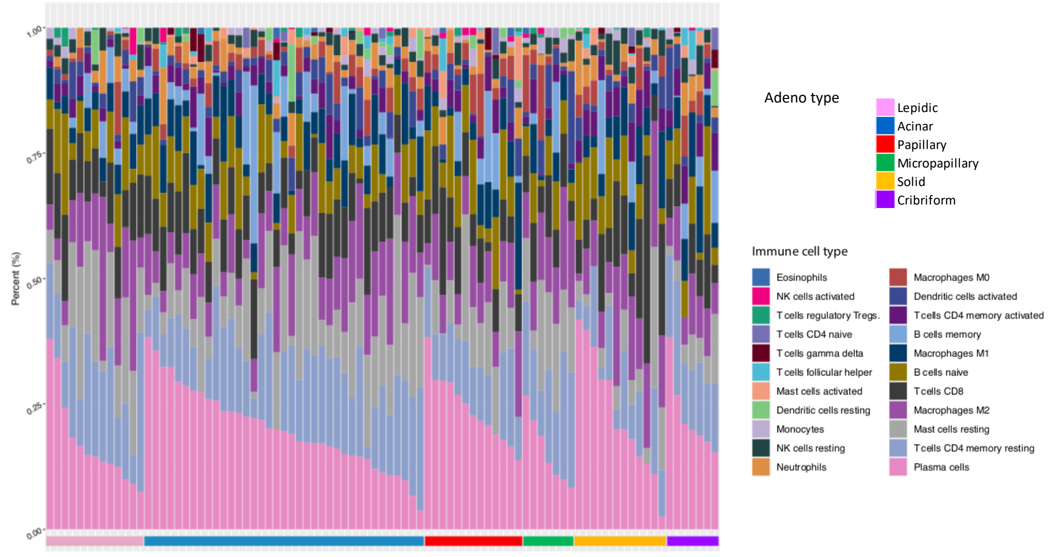


B

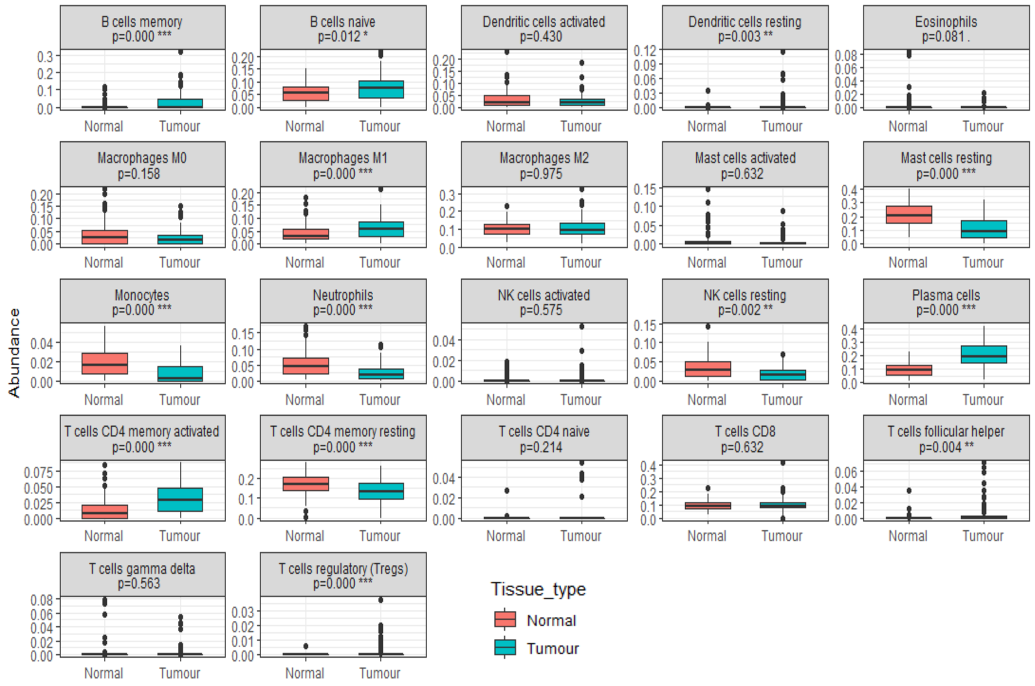


C

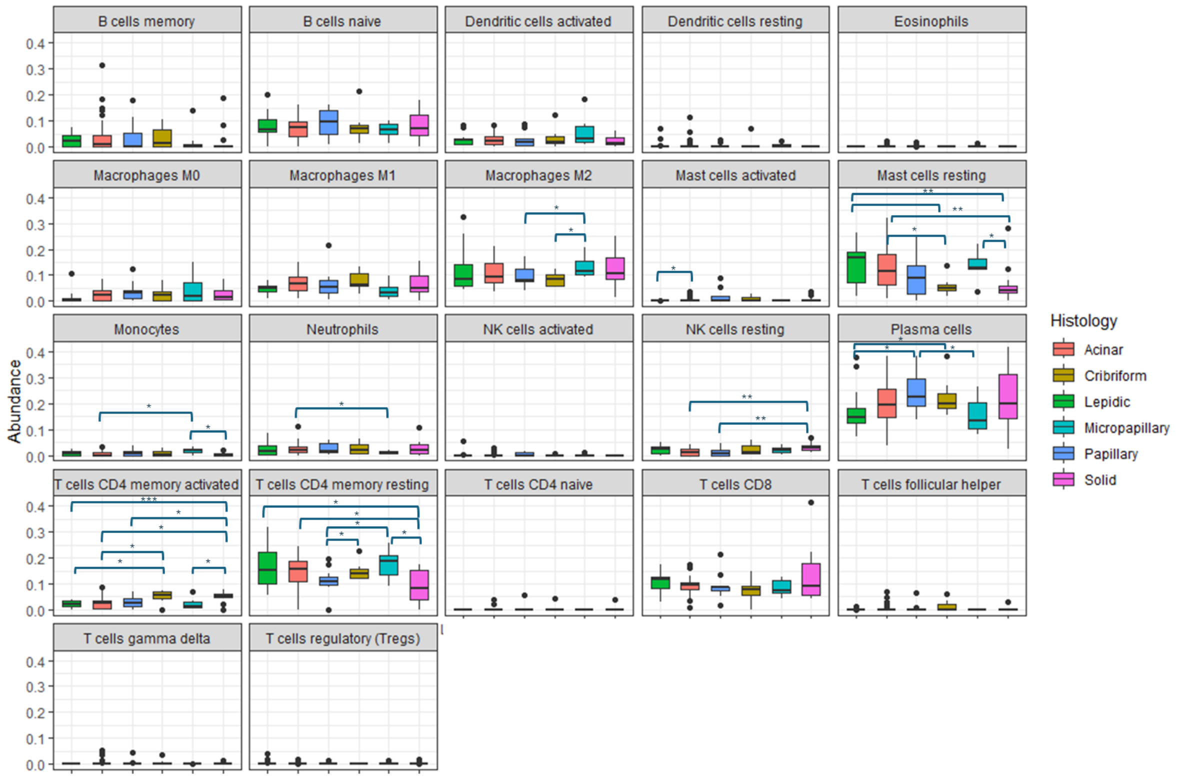


A) Distribution of immune cells in each tumour sample; B) Immune cell abundances in normal versus tumour tissue all samples (Wilcoxon tests of significance with FDR corrections); C) Proportion of immune cells in each tumour histological subtype (Mann-Whitney test with FDR corrections); level of significance: * $P \leq 0.05$; ** $P \leq 0.01$; *** $P \leq 0.001$
